# Defective X-gating caused by *de novo* gain-of-function mutations in *KCNK3* underlies a developmental disorder with sleep apnea

**DOI:** 10.1101/2021.08.05.21261490

**Authors:** Janina Sörmann, Marcus Schewe, Peter Proks, Thibault Jouen-Tachoire, Shanlin Rao, Elena B. Riel, Katherine E. Agre, Amber Begtrup, John Dean, Maria Descartes, Jan Fischer, Alice Gardham, Carrie Lahner, Paul R. Mark, Srikanth Muppidi, Pavel N. Pichurin, Joseph Porrmann, Jens Schallner, Kirstin Smith, Volker Straub, Pradeep Vasudevan, Rebecca Willaert, Elisabeth P. Carpenter, Karin E.J. Rödström, Michael G. Hahn, Thomas Müller, Thomas Baukrowitz, Matthew E. Hurles, Caroline F. Wright, Stephen J. Tucker

## Abstract

Sleep apnea is a common disorder that represents a global public health burden. *KCNK3* encodes TASK-1, a K^+^ channel implicated in the control of breathing, but its link with sleep apnea remains poorly understood. Here we describe a novel developmental disorder with sleep apnea caused by rare *de novo* gain-of-function mutations in *KCNK3*. The mutations cluster around the ‘X-gate’, a gating motif which controls channel opening, and produce overactive channels that no longer respond to inhibition by G-protein coupled receptor pathways, but which can be inhibited by several clinically relevant drugs. These findings demonstrate a clear role for TASK-1 in sleep apnea and identify possible therapeutic strategies.

## Introduction

Sleep apnea is thought to affect up to 1 billion people worldwide and is characterized by abnormal, interrupted breathing during sleep^1,2^. The poor quality of sleep that arises results in huge economic and societal impact, a decreased quality of life, and increases the risk of comorbidities such as cardiovascular disease, diabetes and depression, as well as the risk of motor vehicle accidents^3^. Sleep apnea is therefore a major public health burden and there is a large unmet clinical need for more effective treatments^4^. However, it is also a complex disorder and the mechanisms involved are often unclear.

Increasing evidence suggests that instability of ventilatory control is involved in the pathogenesis of both central and obstructive sleep apnea, the two principal forms of the disorder^1^. Peripheral and central chemoreceptors which detect O_2_/CO_2_ levels also play a critical role, though their molecular identity and the neuronal networks involved remain poorly defined^5^.

Two-Pore Domain K^+^ channels (K2P) are a structurally distinct subset of K^+^ channels where each gene encodes a subunit with two pore-forming domains that co-assemble as a ‘dimer of dimers’ to create a single pseudotetrameric K^+^ selective channel across the membrane^6,7^. K2P channels underlie the background K^+^ currents that control the membrane potential in many different cell types. Originally described as ‘leak’ channels, their activity is now known to be regulated by diverse stimuli, including many G-protein coupled receptor (GPCR) pathways, thus enabling them to integrate different neuronal, metabolic and cellular signaling pathways into changes in cellular electrical activity^8,9^. Such regulatory pathways are known to be important in many of the neural systems that regulate breathing^10^, including the control of respiratory drive immediately after birth where dysfunctional regulation of this pathway gives rise to an increased frequency of spontaneous apneas^11^.

*KCNK3* encodes the TASK-1 K2P channel and is expressed in a variety of neuronal populations throughout the central nervous system, including many chemosensitive regions involved in the control of ventilation as well in hypoglossal and spinal cord motor neurons^12-14^. In peripheral tissues, TASK-1 is also found in the carotid bodies, lung, heart, and pulmonary arterial smooth muscle^15-17^. Its expression throughout these tissues has therefore implicated TASK-1 in the control of breathing and also in sleep apnea^12,18-22^. Furthermore, an X-ray crystal structure of the TASK-1 channel has recently been reported in complex with a compound class of TASK-1 inhibitors currently in clinical trials for the treatment of sleep apnea^23^. This structure also revealed several unique features of TASK-1, including a lower ‘X-gate’, a structural motif which controls opening and closing of the channel pore^23^.

However, a clear mechanistic link between TASK-1 and sleep apnea remains unproven and is further complicated by the propensity of TASK-1 subunits to co-assemble with related TASK-3 (*KCNK9*) subunits to form novel heteromeric TASK-1/TASK-3 channels in cells where both genes are coexpressed^24,25^. Moreover, heterozygous loss-of-function variants in *KCNK3* are associated with a different disorder, pulmonary arterial hypertension (PAH), an adult-onset, progressive and often fatal disease characterized by increased pulmonary arterial pressure in the absence of the common causes of pulmonary hypertension^26^; in addition loss-of-function mutations in the TASK-3 channel (*KCNK9*) are associated with a neurodevelopmental disorder, Birk Barel syndrome^27^.

In this study we describe nine probands with *de novo* missense mutations in *KCNK3* who exhibit global developmental delay, hypotonia, and central/obstructive sleep apnea with a range of structural malformations. The mutations all cluster near the recently identified lower X-gate of the TASK-1 channel and result in a novel gain-of-function phenotype that correlates with the severity of their disorder. These results have important implications for the treatment of these probands and other individuals with sleep apnea, as well as for our understanding of the role that TASK-1 channels play in cellular function.

## Results

### Recurrent missense mutations in KCNK3 cause developmental delay with sleep apnea

A recent analysis of 31,058 parent-offspring trios with severe developmental disorders identified 28 novel disease-causing genes with a high burden of *de novo* mutations, including one caused by recurrent missense variants in *KCNK3* (ENST00000302909; NM_002246)^28^. Through parent-offspring exome sequencing performed across four different diagnostic laboratories and research studies, we have now identified a total of nine probands, each heterozygous for one of six *de novo* missense variants in *KCNK3* (**Methods** and **Table 1**). These novel mutations were found to cluster in two regions of the protein (ENSP00000306275.3; NP_002237): L122V, L122P, G129D (two probands), and N133S (three probands) in the second transmembrane helix (M2); whilst L239P and L241F are in the fourth transmembrane helix (M4) (**Fig. 1a,b**).

**Table 1.** Summary of *de novo* missense mutations in *KCNK3* with proband phenotypes.

| Position (GRCh37) | chr2:26950615 | chr2:26950616 | chr2:26950637 | chr2:26950637 | chr2:26950649 | chr2:26950649 | chr2:26950649 | chr2:26950967 | chr2:26950972 |
| --- | --- | --- | --- | --- | --- | --- | --- | --- | --- |
| HGVSc (NM_002246; ENST00000302909) | c.364C>G | c.365T>C | c.386G>A | c.386G>A | c.398A>G | c.398A>G | c.398A>G | c.716T>C | c.721C>T |
| HGVSp (NP_002237; ENSP00000306275) | p.Leu122Val<br>(L122V) | p.Leu122Pro<br>(L122P) | p.Gly129Asp<br>(G129D) | p.Gly129Asp<br>(G129D) | p.Asn133Ser<br>(N133S) | p.Asn133Ser<br>(N133S) | p.Asn133Ser<br>(N133S) | p.Leu239Pro<br>(L239P) | p.Leu241Phe<br>(L241F) |
| Inheritance | *De novo* | *De novo* | *De novo* | *De novo* | *De novo* | *De novo* | *De novo* | *De novo* | *De novo* |
| Proband gender | Male | Male | Male | Male | Male | Male | Female | Female | Female |
| Age | 16-20yrs | 1-5yrs | (21 weeks) | 6-10yrs | 6-10yrs | 6-10yrs | 6-10yrs | 1-5yrs | 20-25yrs |
| Sleep apnea | Central | Central | NA | Central | Central | Central | Obstructive | Obstructive | Central/Obstructive |
| Nocturnal O2 | Yes | Yes | NA | PEEP | Yes | CPAP | BiPAP | Yes | BiPAP |
| Developmental delay | Mild | Severe | NA | Severe | Moderate | Moderate | Mild | Mild | Mild |
| Learning difficulties | ASD, learning difficulties, speech delay | Speech and language delay, absent speech | NA | Absent speech | ADHD, learning difficulties, speech delay | - | Speech and language delay | Speech and language delay | Speech and language delay, mildly dysarthric |
| Abnormality of musculoskeletal system | Hypotonia, fatiguability, mild scoliosis, torticollis | Hypotonia | Flexion contractures, axillary and perineal pterygia | Hypotonia, arthrogryposis with joint pterygia, severe scoliosis | Hypotonia | Hypotonia, joint hypermobility | Hypotonia, fatigability | Hypotonia | Hypotonia |
| Abnormality of limbs | Talipes calcaneovalgus | Bilateral foot deformities, | Rocker bottom foot | Bilateral talipes equinovarus | Bilateral talipes equinovarus. | Bilateral talipes | Bilateral talipes | - | - |
|  |  | progressive<br>cavovarus |  |  | Left foot fixed,<br>right positional. |  |  |  |  |
| **Abnormality of male**<br>**genitalia/groin** | - | Bilateral<br>undescended<br>testes | Ambiguous<br>genitalia, penile<br>hypospadias | Undescended<br>testes, scrotal<br>hypoplasia | Undescended<br>testes<br>(corrected) | Undescended<br>testes<br>(corrected) | NA | NA | NA |
| **Abnormality of**<br>**digestive system** | Diarrhea,<br>constipation,<br>poor feeder | Constipation | NA | Dysphagia,<br>gastroesophag<br>eal reflux | Constipation,<br>feeding<br>difficulties | - | Constipation | Constipation,<br>dysphagia | Constipation,<br>dysphagia |
| **Abnormality of**<br>**face/jaw** | Facial<br>asymmetry,<br>High arched<br>palate | Wide set eyes,<br>upturned nose,<br>thin upper lip,<br>myopathic<br>facies | Hypertelorism,<br>Micrognathia,<br>Cleft palate,<br>Anteverted<br>nares | Bilateral cleft<br>palate | Deeply set<br>eyes, wide<br>mouth | Tented upper<br>lip vermilion,<br>myopathic<br>facies | Facial<br>weakness,<br>hypoplastic<br>tooth enamel | Facial<br>weakness,<br>tented upper<br>lip, high arched<br>brows | Drooping of both<br>eyelids, ling narrow<br>face |
| **Pregnancy and birth** | - | IUGR,<br>NICU (85 days)<br>polyhydramnios | IUGR,<br>termination of<br>pregnancy | C-section,<br>IUGR,<br>polyhydramnios | - | - | C-section,<br>polyhydramnios | C-section,<br>NICU (8 days) | C-section, NICU (7<br>days) |
| **Weight (current)** | -1.79 SD | +1.37 SD | <-2 SD | -3.68 SD | -0.25SD | -0.63 SD | -0.95 SD | +0.25 SD | -5.07 SD |
| **Height (current)** | -2.27 SD | +0.37 SD | NA | -2.28 SD | +0.42 SD | Unknown | +0.58 SD | -0.35 SD | +0.20 SD |
| **OFC (current)** | -2.20 SD | No recent | <-2 SD | -3.08 SD | -1.55 SD | -1.07 SD | -0.61 SD | -0.10 SD | No recent |

**Acronyms:** PEEP= Positive End Expiratory Pressure; CPAP = Continuous Positive Airway Pressure; BiPAP = Bilevel Positive Airway Pressure; OFC = Occipital Frontal Circumference; NICU
= Neonatal Intensive Care Unit; ASD = Autism Spectrum Disorder; ADHD = Attention Deficit Hyperactivity Disorder; C-section = Caesarean section; IUGR = IntraUterine Growth Restriction;
SD = Standard Deviation

**Figure 1.**
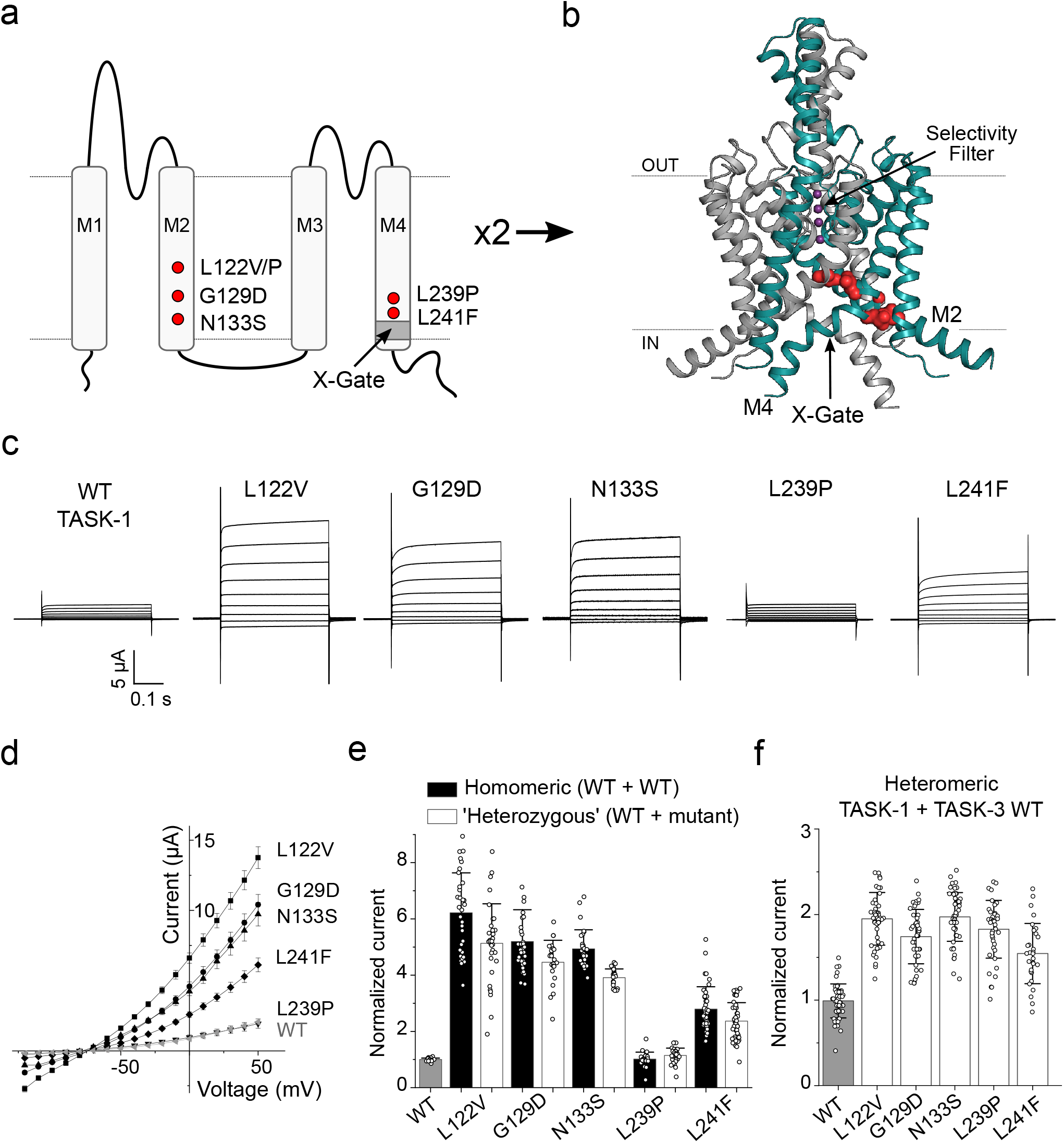
DDSA mutations produce a gain-of-function phenotype in TASK-1. **a)** Topological model of a TASK-1 subunit with position of the DDSA mutants labeled in red and the X-gate in dark grey. Two subunits coassemble to form the K^+^ channel pore **b)** Model showing the dimeric structure of TASK-1 (PDB: 6RV2), with one subunit shown in teal and the residues mutated in DDSA shown as red spheres. **c)** Representative TEVC recordings of WT TASK-1 and DDSA mutant currents in response to a voltage steps from -110 to +50 mV in 20 mV steps from a holding potential of -80 mV. **d)** Current-voltage plot of WT TASK-1 (n = 38), L122V (n=39), G129D (n=44), N133S (n=55), L239P (n=57), and L241F (n=42), data are presented as mean ± S.D. **e)** Currents for homomeric DDSA mutants, and ‘heterozygous’ channels formed from 1:1 coexpression of WT TASK-1 and DDSA mutant normalized to WT currents at +50 mV: TASK-1 (n=28), L122V (n=32), L122V/WT (n=32), G129D (n=36), G129D/WT (n=23), N133S (n=29), N133S/WT (n=23), L239P (n=25), L239P/WT (n=34), L241F (n=42),and L241F/WT (n=43), data are presented as mean ± S.D. **f)** WT or DDSA mutant TASK-1 WT coexpressed 1:1 with WT TASK-3. Currents normalized to WT heteromeric TASK-1/TASK-3 currents: WT (n = 50), L122V (n=49), G129D (n=44), N133S (n=53), L239P (n=41), L241F (n=30), data are presented as mean ± S.D.

All nine probands share similar phenotypes that define this new rare monogenic channelopathy, which we henceforth refer to as ‘Developmental Delay with Sleep Apnea’ (DDSA) (**Table 1**). All living probands (8/9, aged 3-25 years) had hypotonia, global developmental delay, central and/or obstructive sleep apnea that required nocturnal O_2_, and feeding difficulties that necessitated the insertion of a percutaneous endoscopic gastrostomy (PEG) tube. One proband was terminated at 21 weeks due to abnormalities detected during routine ultrasound scanning. The majority of probands (7/9) had major structural malformations, including microcephaly, arthrogryposis/flexion contractures, scoliosis, cleft palate and bilateral talipes, with some facial dysmorphology, and the male probands (5/6) had ambiguous genitalia.

However, two probands (aged 1-5 and 20-25 years) are notably less severely affected, with feeding difficulties, hypotonia and sleep apnea, but only mild developmental delay and no structural abnormalities. Intriguingly, both mutations (L239P, L241F) in these two phenotypically milder individuals are located in the M4 helix of the TASK-1 protein, whereas those in the more severely affected probands (L122V/P, G129D, N133S) are all found in the M2 helix.

### DDSA mutations cluster around the X-gate in TASK-1

Most K2P channels are thought to open and close by controlling a ‘gate’ within their K^+^ selectivity filter^29-31^. However, a recent structure of TASK-1 revealed a novel lower gate formed by an interaction of the pore-lining M4 helices that occludes the entrance to the pore^23^. Importantly, this closed ‘X-gate’ structure contains a six amino acid motif (VLRFMT) previously implicated in the control of channel activity by volatile anesthetics and GPCRs, though the mechanisms involved in opening and closing the pore remain unclear^32,33^.

Upon examination of the TASK-1 crystal structure (PDB ID: 6RV2), we found that, even though these six DDSA mutations are on two different transmembrane helices (M2 and M4), they all cluster near the X-gate and are involved in either inter- and/or intra-subunit interactions likely to hold the X-gate closed (**Fig. 1b** and **Supplementary Fig.1a,b**). Given that several of the *de novo* mutations are recurrent, and that loss-of-function variants in *KCNK3* are associated with a completely different adult-onset disorder^26^, we hypothesized that DDSA may be caused by a different mechanism.

### DDSA mutations produce a gain-of-function phenotype in TASK-1

We therefore examined the functional activity of all these DDSA mutations by heterologous expression of either wild-type (WT) or mutant channels in *Xenopus* oocytes. Two-Electrode Voltage Clamp (TEVC) recordings revealed whole-cell K^+^ currents markedly larger than WT TASK-1. Only one variant, L239P in the M4 helix, produced whole-cell currents similar in size to WT TASK-1 (**Fig. 1c,d** and **Supplementary Fig. 2a**).

The mutant currents all exhibited reversal potentials around -80 mV in low external [K^+^] consistent with K^+^ selective channels (**Fig. 1d**). The L122V mutation produced the largest increase in current compared to WT consistent with the activatory effect of mutations at the structurally equivalent position in many other K2P channels^34^. Overall, the mutations located in M2 produced the largest increase in current compared to WT TASK-1 (3.2-6.2-fold), whereas the two M4 mutations were either similar in size to WT (i.e. L239P), or only ∼2.8-fold (i.e. L241F; **Fig. 1d**).

### Gain-of-function in ‘heterozygous’ TASK-1 and heteromeric TASK-1/TASK-3 channels

K2P channels are dimeric and the results described above were all obtained in homomeric channels where both subunits contain the mutation. However, the DDSA probands are heterozygous for a single mutation, meaning a mixture of channels can be created from coassembly of WT and mutant subunits (25% WT, 50% ‘heterozygous’ and 25% ‘homomeric’ mutant). Therefore to better replicate this heterozygous genotype, we co-injected oocytes with WT TASK-1 mRNA plus an equal amount of either WT or mutant mRNA in a 1:1 ratio. In all cases, the resulting currents were at least 80% the size of currents formed by homomeric mutant channels (**Fig. 1e)**. An increase in current was also observed for the most common DDSA variant (N133S) in a ‘pure’ heteromeric channel formed from a covalently linked WT/mutant tandem dimer which constrains channel stoichiometry (**Supplementary Fig. 2b**).

TASK-1 can also coassemble with TASK-3 subunits to form novel heteromeric channels *in vivo*^24,25^. We therefore co-injected oocytes with WT TASK-3 mRNA plus an equal amount of either WT or mutant TASK-1 mRNA. In this case, all the mutants, including L239P, produced larger heteromeric TASK-1/TASK-3 currents than when WT TASK-1 was used (**Fig. 1f**). This demonstrates that all these DDSA mutations produce a gain-of-function in either heterozygous TASK-1 and/or heteromeric TASK-1/TASK-3 channels. This effect is clearly different to the loss-of-function observed for *KCNK3* mutations associated with PAH^26^, including the recently characterized missense variant, L214R^35^; **Supplementary Fig. 2c,d**). In further support of our findings, we did not observe any functional effect of a control variant (H141Q) (**Supplementary Fig. 2e**). This variant, which is also located in the M2 helix, is present in 194 individuals in the gnomAD database^36^ and annotated as likely benign in ClinVar^37^.

### DDSA mutations increase channel open probability

The whole-cell currents (*I*) described above are a product of the number of channels in the membrane (*N*) and their individual open probability (*P*_*o*_). However, trafficking of mutant proteins to the membrane is normally impaired by mutations and rarely increased; also membrane trafficking in an oocyte does not necessarily reflect what happens in more complex native cells, especially neurons^17,38^. We therefore measured the properties of single TASK-1 channels for the N133S variant in M2, and for L239P in M4 (**Fig. 2a-d**). Similar to previous reports, we found the *P*_*o*_ of WT TASK-1 was extremely low (*P*_*o*_ ∼ 0.02) with a single channel conductance of ∼13 pS^39^. However, homomeric N133S mutant channels exhibited an increased *P*_*o*_ ∼10-fold greater than WT TASK-1 (**Fig. 2c**) with an unchanged single channel conductance (**Fig. 2d**), an effect consistent with previous reports of other mutations at this position in both TASK-3 and TASK-1^23,40^. We also observed a similar effect for the L239P mutation with a 10-fold increase in estimated *P*_*o*_, though its “blurry” open channel histogram indicates that its effects on single channel properties are more complex and warrant further investigation (**Fig. 2b**).

**Figure 2.**
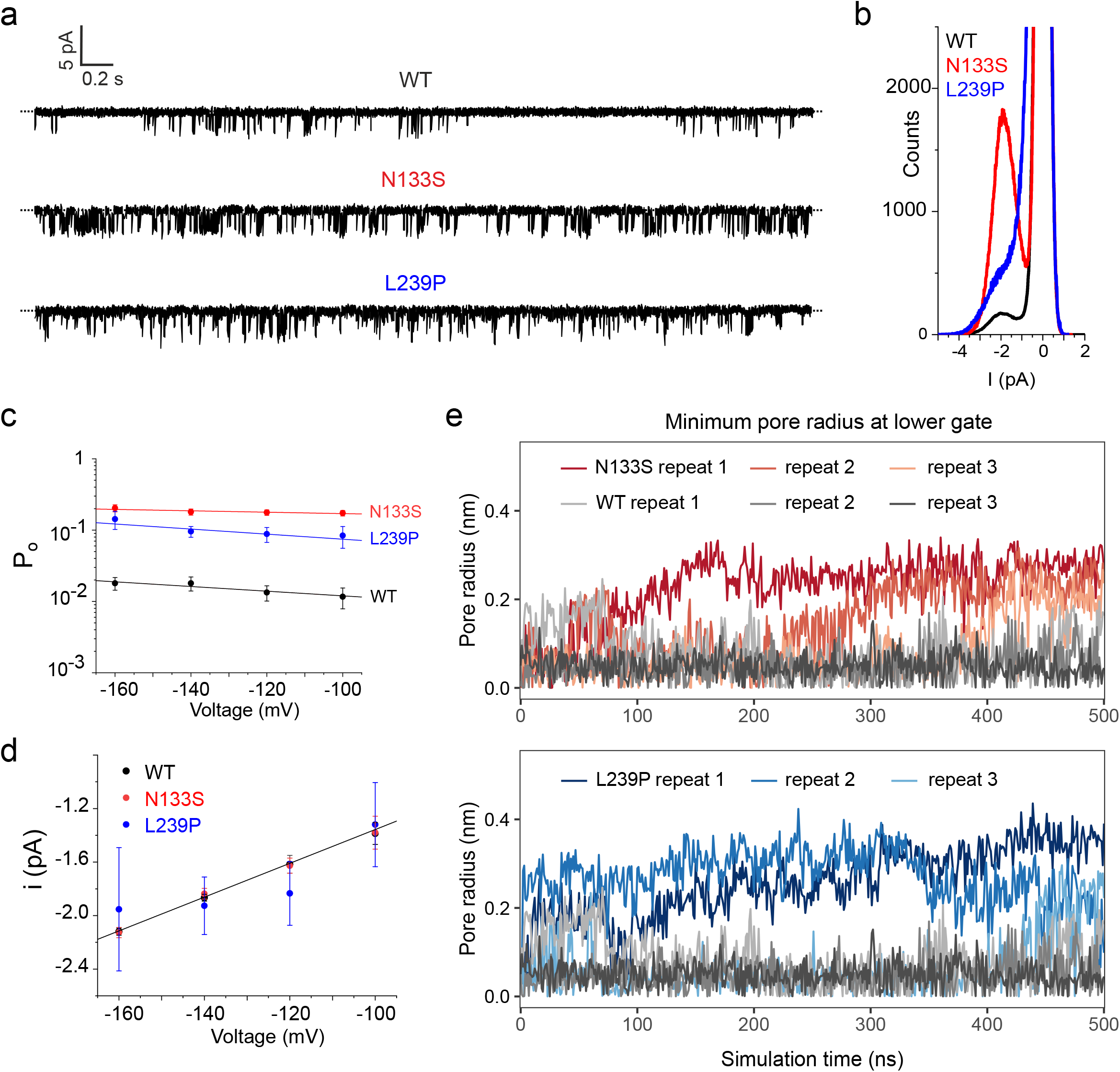
Increased channel open probability due to destabilization of the X-gate. **a**) Representative single channel recordings at a holding potential of -160 mV. **b**) Corresponding histogram normalized to a zero-current level (closed state). **c**) Single-channel open probability (*P*_*o*_) and **d**) single channel conductance at different potentials for TASK-1 WT (n=3), N133S (n=3), L239P (n=4), data are presented as mean ± S.D. A straight-line fit is shown for the WT single channel conductance values only **e,f**) Plot of the minimum pore radius at lower X-gate during three independent repeats of molecular dynamics simulations of the WT TASK-1 structure compared to either the N133S mutant structure, and the L239P mutant.

### DDSA mutations destabilize the closed X-gate in TASK-1

Destabilization of the interactions which hold the channel closed will increase the frequency of channel openings (i.e. increase *P*_*o*_) and is consistent with the location of these DDSA mutations near the X-gate. In particular, the recurrent mutation N133S on M2 involves changes to an amino acid that participates in a hydrogen bond predicted to stabilize the X-gate when closed^23,40^. To investigate this further, we examined multiple substitutions at this position, all of which resulted in a gain-of-function indicating the critical role of this hydrogen bond (**Supplementary Fig. S3a,b**).

In addition, we performed molecular dynamics simulations on WT and both N133S and L239P mutant channel structures. These structures were embedded within a lipid bilayer and multiple repeats of equilibrium simulations each run for 500 ns. The results show the lower X-gate remained firmly closed in all simulations of the WT channel, but began to open in simulations of the N133S and L239P mutant channels (**Fig. 2e**). Interestingly, we previously proposed a twist and straightening was required to open the X-gate^23^ and similar movements were observed in simulations of the N133S mutant channel (**Supplementary Fig. S3c**).

### DDSA mutations result in defective X-gating and dysfunctional GPCR-mediated inhibition

TASK channels have been shown be inhibited *in vivo* by multiple hormones and transmitters including ATP, thyrotropin-releasing hormone, serotonin, glutamate, catecholamines and acetylcholine that all act through G protein-coupled receptors (GPCR) signaling via G proteins of the Gαq/11 subclass (Gαq)^9,13,17,32^. Furthermore, mutation of residues in M2 and in the X-gate itself have been shown to impair this process^32,33^. We therefore examined the ability of these DDSA mutations to affect such regulation.

To reflect the heterozygous probands, we co-expressed equal amounts of WT and mutant TASK-1 mRNAs and measured their inhibition by either carbachol via endogenous muscarinic receptors, or by ATP via a coexpressed P2Y receptor as reported previously^41,42^. However, unlike WT TASK-1, we found that GPCR-mediated inhibition by either approach was markedly impaired in all ‘heterozygous’ DDSA mutant co-expressions (**Fig. 3a-d**). This effect was similar to that observed when these mutations were expressed as homomeric channels (**Supplementary Fig. S4a**). By contrast, GPCR-regulation of the ‘benign’ H141Q variant appeared unaffected (**Supplementary Fig. S4b**).

**Figure 3.**
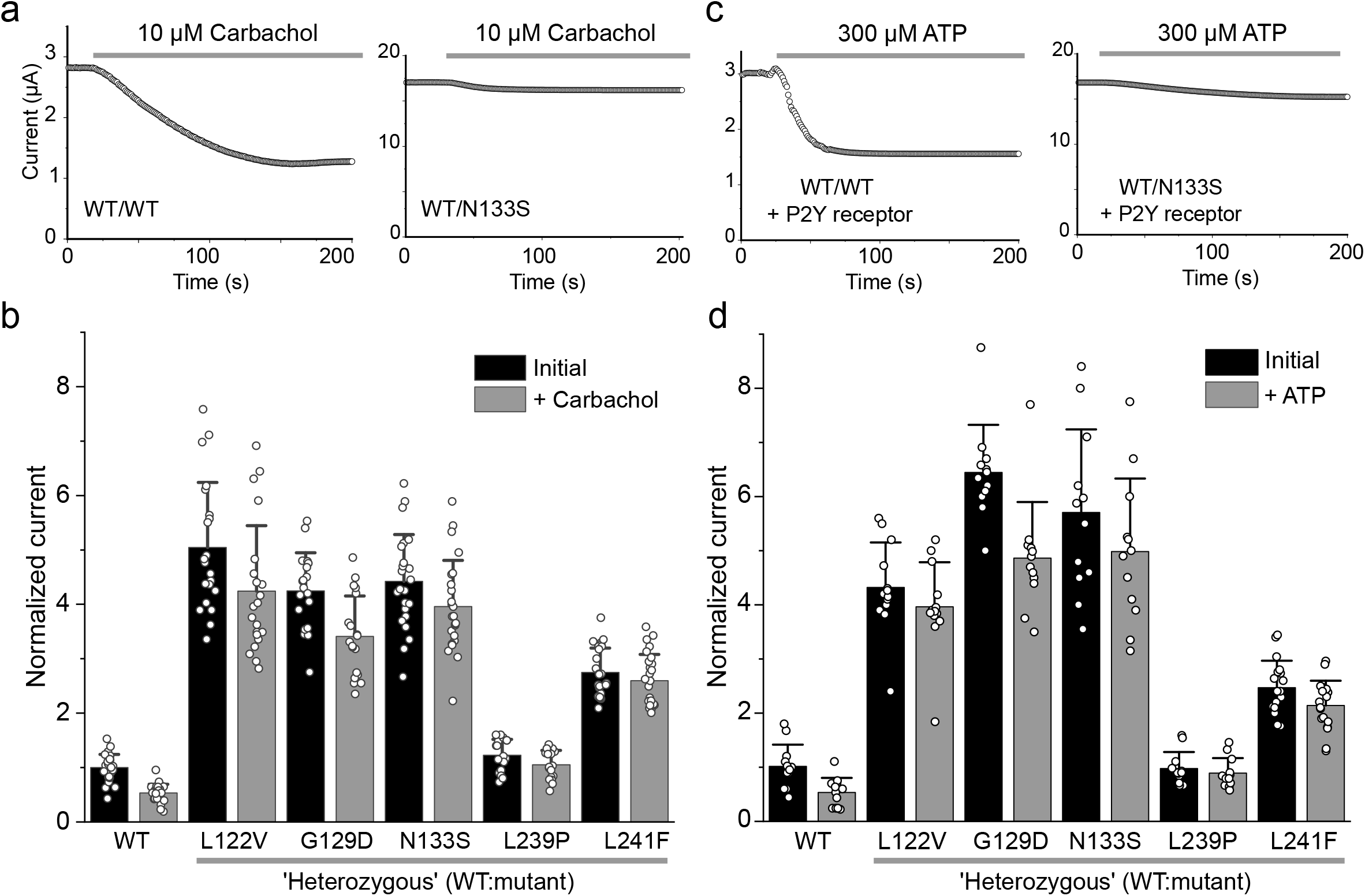
Dysfunctional GPCR-mediated inhibition in DDSA mutants. **a)** Representative currents at +50 mV of TASK-1 WT and coexpressed WT/N133S over time while adding 10 µM Carbachol which elicits ∼50% inhibition of WT TASK-1. **b)** Currents normalized to initial WT current for WT TASK-1 coexpressed 1:1 with the indicated DDSA mutant before and after addition of 10 µM carbachol. WT TASK-1 (n = 26), L122V (n=21), G129D (n=19), N133S (n=24), L239P (n=18), L241F (n=26), data are presented as mean ± S.D. **c,d)** Equivalent recordings for WT TASK-1 coexpressed 1:1 with each DDSA mutant as indicated and the P2Y receptor (1:1:4). The current levels shown are before and after addition of 300 µM ATP normalized to the initial WT current. TASK-1 (n = 12), L122V (n=13), G129D (n=12), N133S (n=12), L239P (n=12), L241F (n=18), data are presented as mean ± SD.

The impaired GPCR-mediated inhibition of these DDSA mutations therefore amplifies their gain-of-function effect such that, after GPCR activation, their remaining currents were all greater than WT TASK-1. Importantly, this effect was notable even for the WT/L239P channel, which was initially similar in size to WT TASK-1, but the currents were ∼2-fold larger than WT after receptor stimulation (**Fig. 3b,d**). This GPCR-insensitivity also affected heteromeric TASK-1/TASK-3 channels that contained mutant TASK-1 subunits (**Supplementary Fig. S4c**).

### Druggability of DDSA mutations

Unlike many other K2P channels which exhibit relatively poor pharmacology^31^, TASK-1 can be inhibited by a range of potent and highly selective small molecules, including a number of clinically relevant drugs such as doxapram, carvedilol and bupivacaine^43^. The most potent inhibitor of TASK-1 known to date is BAY1000493 which binds deep within the pore of the channel^23^, and is a close analog of another TASK-1 inhibitor recently used in a clinical trial for the treatment of obstructive sleep apnea (the ‘KOALA’ trial, NCT04236440 currently being progressed into a larger Phase 2 study).

Using our ability to record TASK-1 activity in giant excised membrane patches, we were able to accurately measure the inhibition of WT TASK-1 by BAY1000493 (**Fig 4a,b**); this produced half-maximal inhibition at a concentration of ∼1 nM (IC_50_= 970 pM ± 250 pM, n=10) making it the most potent TASK-1 inhibitor known to date. 1 nM BAY100493 was slightly less effective on TASK-1 N133S and L241F mutant channels (**Fig. 4a**), but full dose-response measurements revealed that all five DDSA mutants still exhibited sensitivity to this drug within the low nanomolar range with IC_50_ values ranging from 5 - 23 nM (**Fig. 4b**). We also examined a range of other known high affinity inhibitors of WT TASK-1 on the N133S mutation; these included PK-THPP, A1899, A293 and Tetrapentylammonium (TPA)^43^, all of which we found to have similar IC_50_ values compared to WT TASK-1 (**Fig. 4c**). This mutation also did not affect the reported sensitivity of TASK-1 to inhibition by several drugs already in clinical use including bupivacaine, carvedilol, and the respiratory stimulant doxapram (**Fig 4c**). This result is in marked contrast to the reported effect of other gain-of-function mutations, e.g. the L244A and M247A mutations engineered into the X-gate of TASK-1 which also both increases channel activity, but which markedly reduce BAY1000493 sensitivity^23^. The druggability of these DDSA gain-of-function mutations therefore highlights potential therapeutic strategies for these probands.

**Figure 4.**
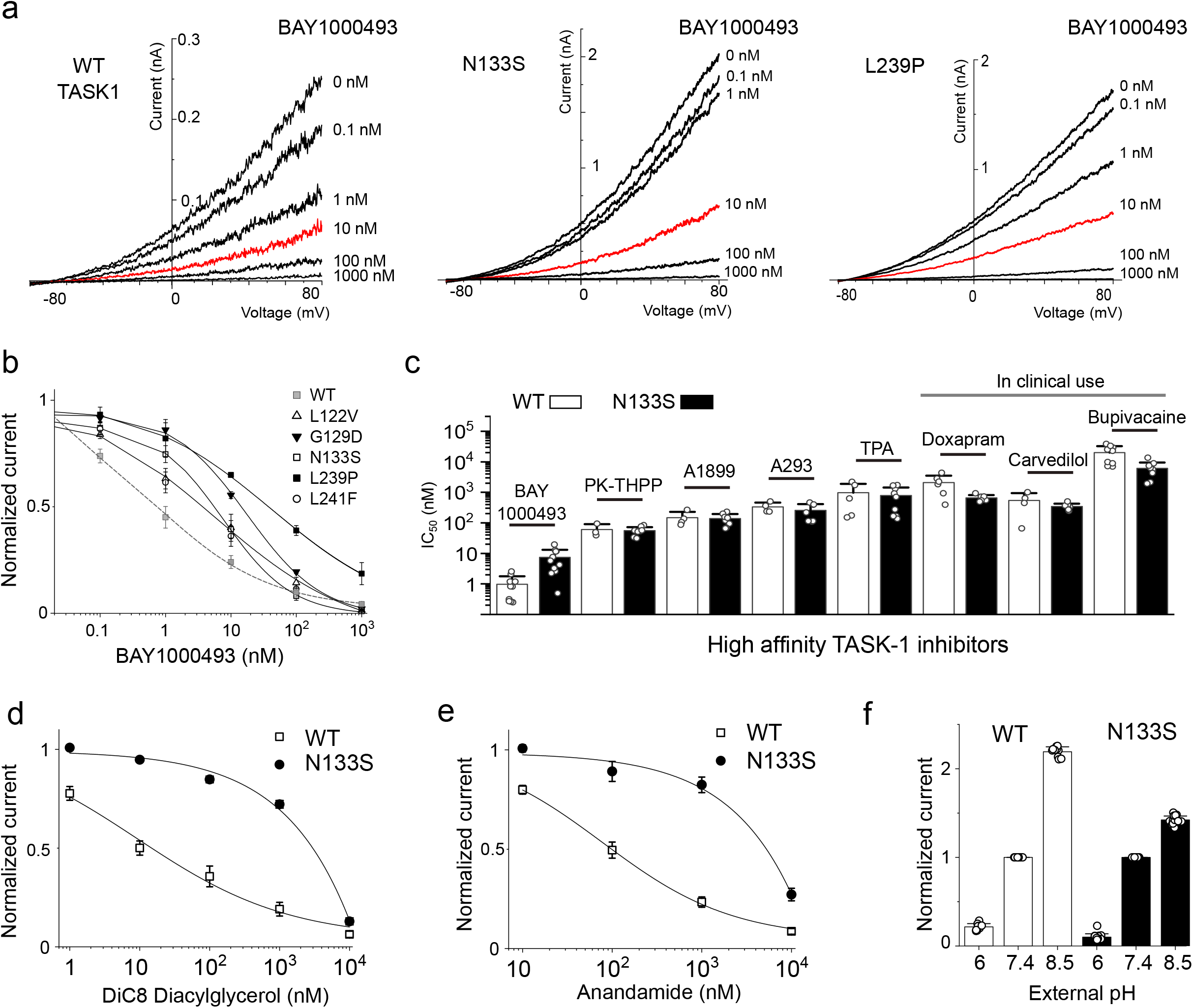
Mechanism of regulation and pharmacology. **a)** Representative excised membrane patch recordings of either WT TASK-1, N133S or the L241F mutant channel activity in response to different concentrations of BAY1000493 applied to the intracellular side of the patch (10 nM shown in red). **b)** Corresponding dose response curves for inhibition of either TASK-1 or DDSA mutants by BAY1000493; WT TASK-1 (n = 8), L122V (n=10), G129D (n=10), N133S (n=11), L239P (n=3), L241F (n=10), data are presented as mean ± S.E.M. Values for WT TASK-1 fitted with grey dashed line. **c)** Comparison of IC_50_ values of various high affinity TASK-1 inhibitors on either WT TASK-1, or the N133S mutant. For BAY1000493: WT (n=10), N133S (n=11); PK-THPP: WT (n=3), N133S (n=9); A1899: WT (n=4), N133S (n=9), WT (n=4), A239: N133S (n=7); TPA: WT (n=5), N133S (n=9); Doxapram: WT (n=6), N133S (n=6); Carvedilol: WT (n=4), N133S (n=7), Bupivacaine: WT (n=9), N133S (n=10). **d**) Dose-response curves for inhibition of either WT TASK-1 (n=12) or the N133S mutant (n=9) in excised patches by the analog of diacylglycerol, DiC8. **e**) Similar dose response curves for inhibition of WT TASK-1 (n=8) and the N133S mutant (n=9) by anandamide in excised patches. In both cases the data are presented as mean ± S.E.M. **f)** Normalized whole-cell currents for WT TASK-1 (n=10) or N133S mutant (n=15) channels at +50 mV at indicated external pH values normalized to initial currents recorded at pH 7.4.

### Mechanism of GPCR-mediated inhibition

The molecular mechanism of Gαq-mediated inhibition is unclear and reported to involve non-canonical effects of pathway activation^41,44^. Normally, Gαq activates phospholipase C (PLC) which hydrolyses phosphatidylinositol (4,5) bisphosphate (PIP_2_), a lipid which supports the activity of many K2P channels, and so PIP_2_ degradation usually reduces channel activity^9,41,45^. However, it appears that it is the concomitant increase in diacylglycerol (DAG) upon PIP_2_ hydrolysis which is primarily responsible for TASK channel inhibition rather than the loss of PIP_2_^41,44-46^. DAG has been shown to directly inhibit TASK-3 channel activity^44^ and we now show that the DAG analogue, DiC8 also directly inhibits TASK-1 with nanomolar efficacy in excised patches. However, this inhibitory effect is severely impaired by the recurrent DDSA mutation, N133S (**Fig. 4d**).

PLC activation can also affect the levels of many other signaling lipids, including the endocannabinoid, anandamide (AEA) which directly inhibits TASK-1^47^, and we found that the N133S mutation also impairs the inhibitory effect of AEA (**Fig. 4e**). Extracellular pH is also known to regulate TASK channel activity^7^. We therefore examined the response of the N133S mutation to changes in extracellular pH and found that although its activation by alkalinization was blunted, its inhibition by extracellular H^+^ remained mostly intact (**Fig. 4f**).

## Discussion

In this study, we describe a new monogenic channelopathy, Developmental Delay with Sleep Apnea (DDSA), resulting from heterozygous *de novo* gain-of-function mutations in *KCNK3*. These mutations result in defective X-gating of TASK-1 and increased K^+^ currents through these channels. Although DDSA is rare, our results also now highlight the important link between TASK-1 channels and sleep apnea, and identify therapeutic opportunities for both DDSA patients and those who suffer from sleep apnea.

TASK-1 channels (including heteromeric TASK-1/TASK-3) have long been regarded as attractive targets for the treatment of sleep apnea^48^. However, even though drugs that target TASK-1 are currently being progressed into Phase 2 clinical trials for treatment of sleep apnea, a clear link with this disorder has not been established and has even been questioned^49^. Furthermore, until now, the only known genetic mutations in TASK-1 were associated with a completely different, hypertensive phenotype^26^.

Our results therefore provide the first direct evidence of a link between *KCNK3* and sleep apnea due to a new class of *de novo* gain-of-function mutations that have very different effects on cellular electrical activity compared to the loss-of-function mutations associated with PAH. This causal link is not only consistent with expression of TASK-1 in many of the chemosensitive cell types and tissues involved in regulation of breathing, but also with the fact that current clinical trials for sleep apnea aim to suppress TASK-1 K^+^ channel activity.

The DDSA probands identified in this study all have a developmental disorder that includes sleep apnea as well as a number of other cognitive and musculoskeletal phenotypes (see **Table 1**). It is well known that inappropriate spatiotemporal expression of an ion channel can affect the development of many cell types, especially neurons^50^ and studies in developing mouse embryos have shown that *KCNK3* is expressed at an early stage especially in the developing CNS^14^. It is also important to note that the effect of these DDSA mutants on electrical activity will be very different to the loss-of function PAH mutations in TASK-1. The phenotype in these probands could therefore arise via several possible mechanisms: a general increase in homomeric TASK-1 activity, increased heteromeric TASK-1/TASK-3 activity, and/or the resistance of these channels to inhibition by Gαq-coupled signaling pathways (see **Fig. 5**). However, the relative contribution of these different molecular mechanisms will be difficult to dissect.

**Figure 5.**
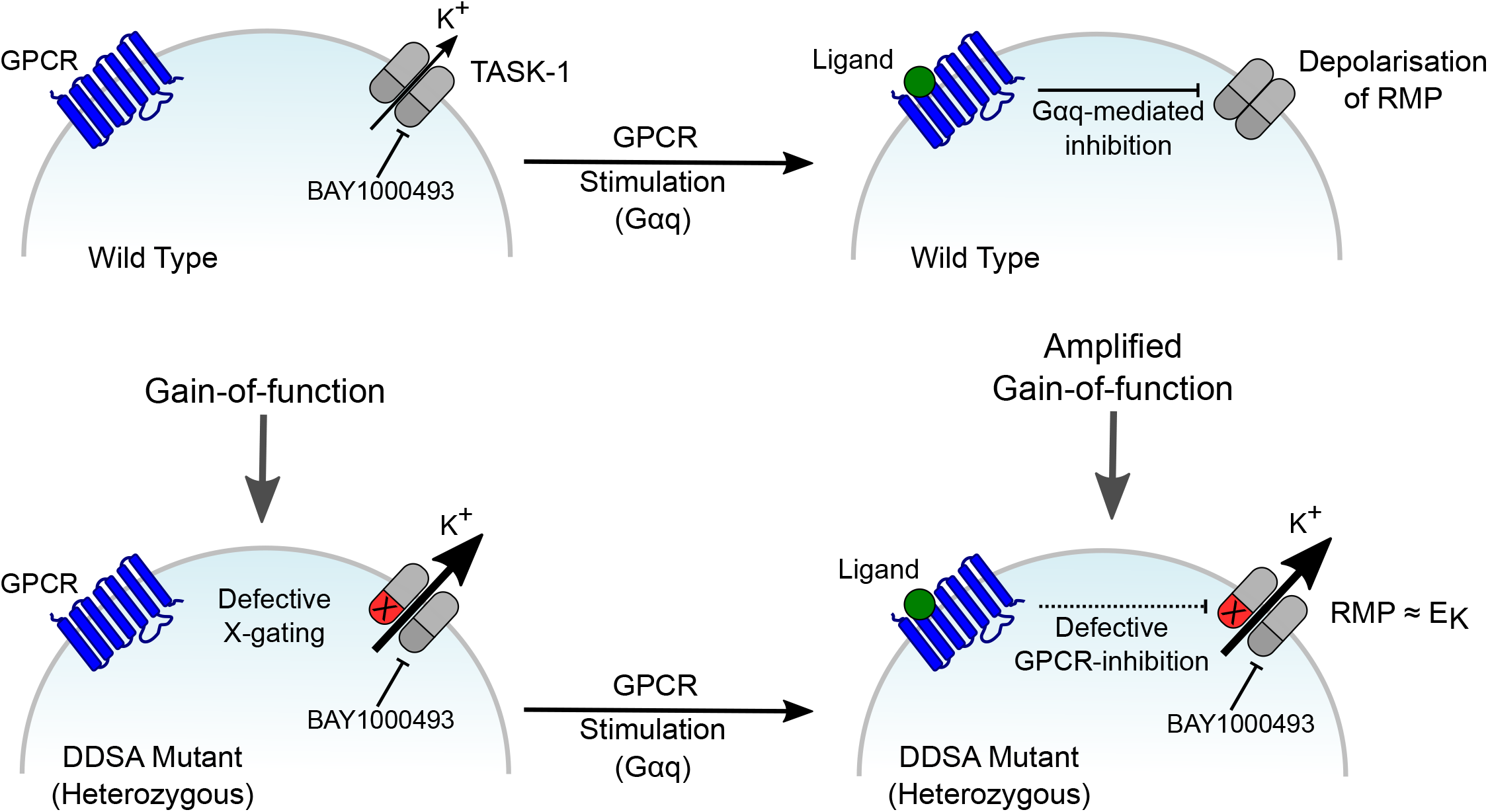
Proposed model for effect of DDSA mutants on cellular electrical activity. In wild type cells the activity of TASK-1 (i.e. homomeric TASK-1 and/or heteromeric TASK-1/TASK3) channels contribute to the hyperpolarized resting membrane potential (RMP). This activity can be inhibited by stimulation of Gαq-coupled receptor pathways and results in depolarization of the RMP. This gating process involves closure of the cytoplasmic X-gate. However, in cells with a single heterozygous DDSA mutation in TASK-1, these mutations (marked as X) result in defective closure of the X-gate (marked in red). Consequently, TASK-1 channel activity is either increased and/or unresponsive to GPCR-mediated inhibition which amplifies the underlying gain-of-function. This increased channel activity keeps cells hyperpolarized near the RMP and also uncouples them from regulation by many different GPCR signaling pathways. Importantly, mutant channels retain sensitivity to inhibition by several high affinity small molecule inhibitors including BAY1000493 that is similar to compounds currently in clinical trial for the treatment of sleep apnea. This offers a range of possible therapeutic strategies for these probands and strengthens their proposed use for the treatment of sleep apnea.

Despite the many phenotypic features shared by the probands, the two with mutations in the M4 helix were much less severely affected than the probands with mutations in the M2 helix, including only mild developmental delay and no structural abnormalities. Interestingly, these M4 mutations (L239P and L241F) each produced the smallest overall increase in whole-cell currents. This genotype-phenotype correlation may therefore provide insight into the mechanisms underlying the DDSA phenotypes, but further studies are required to confirm this. Strikingly, central and/or obstructive sleep apnea was experienced by all (living) probands, and the general gain-of-function in TASK-1 is common to all these variants. We also note that, despite the reported overexpression of *KCNK3* in atrial fibrillation^51^, none of these DDSA probands exhibited any obvious cardiac phenotype.

The functional properties of these mutations provides further insight into the molecular mechanism of TASK-1 gating; in particular, how the X-gate opens and how this is regulated by Gαq-coupled pathways. In the structure of TASK-1, the channel pore is occluded by the lower X-gate^23^. The six residues in M4 that form this constriction were previously identified as important for channel regulation by volatile anesthetics and by several Gαq-coupled pathways, though the mechanisms involved were unclear^32^. Our results show that the DDSA mutations all cluster near the X-gate in its closed conformation and are likely to disrupt stability of the closed X-gate. In particular, the recurrent N133S variant destabilizes a critical H-bond and promotes channel opening. Likewise, the inhibitory effect of DAG on TASK-1 channels suggest that it favors stability of the closed state. The fact that all DDSA mutations produce similar effects in both homomeric and ‘heterozygous’ channels also suggests that disruption of the X-gate within a single subunit is sufficient to open the channel.

The increased whole-cell currents generated by these DDSA mutants result primarily from an increase in channel *P*_*o*_ rather than an increased number of channels at the cell surface, and in some cases the mutants may even decrease surface expression; e.g. the L239P mutant has a 10-fold greater *P*_*o*_, but has similar whole-cell currents to WT TASK-1, an effect that can only result from fewer channels in the membrane. However, Gαq-mediated inhibition of the L239P mutant channel is also completely abolished and so the remaining currents become ∼2-fold larger than WT TASK-1 and therefore represent a gain-of-function effect.

Fortunately, despite their activatory effects, these mutants all retain their sensitivity to a number of small molecule inhibitors, some of which are already in clinical use and/or ongoing clinical trials. This offers a realistic prospect for therapeutic intervention in DDSA probands that may improve their quality of life. It remains to be determined precisely how TASK-1 dysfunction produces the phenotype observed in DDSA, in particular sleep apnea. However, our findings markedly strengthen the proposed use of TASK-1 inhibitors to treat sleep apnea and provide important new insights into a sleep disorder that severely impacts many millions of lives.

## Data Availability

All required data is contained within the paper

## Acknowledgments

This work was supported by grants from the Biotechnology and Biological Sciences Research Council to S.J.T (BB/T002018/1 and BB/S008608/1) and from the Deutsche Forschungsgemeinschaft to M.S. (SCHE 2112/1-2) and T.B (BA 1793/6-2) as part of the Research Unit FOR2518, DynIon The DDD study presents independent research commissioned by the Health Innovation Challenge Fund [HICF-1009-003] a parallel funding partnership between the Wellcome Trust and the Department of Health, and the Wellcome Sanger Institute (WT098051). See www.ddduk.org/access.html for full acknowledgement. S.J.T and E.P.C also received support from the Wellcome Trust as part of the OXION Initiatives in Ion Channels and Membrane Transport in Health and Disease (WT084655MA and 102161/B/13/Z)

## Methods

### Genetics

Four probands were identified through the UK Deciphering Developmental Disorders (DDD) Study, which has been described previously^52,53^. Briefly, probands with severe, undiagnosed developmental disordered and their parents were recruited and phenotyped by referring clinical geneticists in 24 regional genetics services across the National Health Services in the UK and Ireland. Saliva and/or blood-extracted DNA samples were analysed at the Wellcome Sanger Institute using massively parallel whole exome sequencing of the family trio using SureSelect RNA baits (Agilent Human All-Exon V3 Plus with custom ELID C0338371 and Agilent Human All-Exon V5 Plus with custom ELID C0338371). Enriched libraries were analysed by 75-base paired-end sequencing (Illumina HiSeq); reads were mapped to GRCh37/UCSC hg19 and variants called using the Genome Analysis Toolkit (GATK)^54^ and annotated using Ensembl Variant Effect Predictor (VEP)^55^. *DeNovoGear* was used to predict likely *de novo* mutations^56^ and variants were identified for clinical feedback as previously described^57^.

Three probands were identified by the USA-based diagnostics company GeneDX. Using genomic DNA from the proband and parents, the exonic regions and flanking splice junctions of the genome were captured using the Clinical Research Exome kit (Agilent Technologies, Santa Clara, CA) or the IDT xGen Exome Research Panel v1.0. Massively parallel (NextGen) sequencing was done on an Illumina system with 100bp or greater paired-end reads. Reads were aligned to human genome build GRCh37/hg19, and analyzed for sequence variants using a custom-developed analysis tool. Additional sequencing technology and variant interpretation protocol has been previously described^58^. The general assertion criteria for variant classification are publicly available on the GeneDx ClinVar page (http://www.ncbi.nlm.nih.gov/clinvar/submitters/26957/).

One proband was identified by the Mayo Medical Laboratories in the USA. Whole exome sequencing was performed on genomic DNA extracted from all samples submitted. The exome was captured utilizing a custom reagent developed by Mayo Clinic and Agilent Technologies. Sequencing was performed on an Illumina HiSeq 2500 Next Generation sequencing instrument, using HapMap Sample NA12878 as an internal control. Paired-end 101 base-pair reads were aligned to a modified human reference genome (GRCh37/hg19) using Novoalign. Sequencing quality was evaluated using FastQC. All germline variants were jointly called through GATK Haplotype Caller and GenotypeGVCF and each variant was annotated using the BioR Toolkit. Using a custom-developed analysis tool, data were filtered and analyzed to identify clinically relevant sequence variants. Variants of interest were confirmed by automated Sanger sequencing.

One proband was identified at the German Institute of Clinical Genetics Dresden, Germany. DNA was extracted from blood-derived lymphocytes using QIAamp DNA Blood Mini Kit (Qiagen). Patient as well as parental samples were studied in parallel (trio exome strategy). Exome capture was performed using IDT Xgene exome research panel. 150 nucleotide paired-end sequencing was performed with a median target coverage of at least 50-fold on Illumina NextSeq500 Sequencing systems. Alignment (mapping to GRCh37/hg19), variant identification (SNVs and indels), variant annotation and filtering was performed using the CLC Biomedical Genomics Workbench (Qiagen, Hilden, Germany) as described previously^59^. Variants were filtered with a focus on protein-altering variants (missense, frameshift, splice-site and premature stop-codons), rare in public databases (gnomAD, allele frequency below 1% and not more than 5 homozygous or hemizygous individuals). Additionally, variants were prioritized based on assumed inheritance patterns (de novo heterozygous, homozygous, hemizygous and compound heterozygous).

For all studies, candidate *de novo* mutations in *KCNK3* were visually inspected using the Integrative Genomics Viewer (IGV)^60^. Likely diagnoses were communicated to referring clinical teams for diagnostic validation (including confirmation by Sanger sequencing where appropriate) and discussion with the family.

### Electrophysiology

The wild-type human TASK-1 gene (*KCNK3*) was subcloned into a plasmid vector (pFAW) suitable for *in vitro* transcription and expression in *Xenopus laevis* oocytes^61^. Mutations were introduced by site-directed mutagenesis and confirmed by sequencing. Unless otherwise stated, a volume of ∼18 nl of mRNA was injected oocytes at a concentration of 110 ng/μl for either wild-type or mutants (i.e. 2 ng of RNA per oocyte). Two-electrode voltage clamp recordings were performed as previously described^61^. Briefly, after injection of mRNA, oocytes were incubated for 20–24 h at 17.5°C and measured in ND96 buffer at pH 7.4 (96 mM NaCl, 2 mM KCl, 2 mM MgCl2, 1.8 mM CaCl2, 5 mM HEPES). Unless otherwise stated, currents were recorded using a 400 ms voltage step protocol from a holding potential of -80 mV delivered in 10 mV increments between -120 mV and +50 mV and 800 ms ramp protocols from -120 to +50 mV. All recorded traces were analyzed using Clampfit (Axon Instruments), and graphs were plotted using Origin2019b (OriginLab Corporation). Unless otherwise described, all results shown are reported as mean ± standard deviation and obtained with oocytes from at least 3 independent batches.

Single-channel currents in cell-attached patches from *Xenopus* oocytes were recorded in symmetrical 140mM KCl solutions containing 10mM HEPES (pH 7.4 with KOH). Data was filtered at 2 or 5 kHz and recorded at a 200 kHz sampling rate with program Clampex on an Axopatch 200B amplifier. Data analysis was performed using Clampfit. Single channel current amplitude of the L239P mutant channels was estimated as follows. First, a closed level histogram for current values above 0 pA was approximated as a mirror image of the values below 0 pA and the resulting histogram was subtracted from the data. The mean single channel current amplitude *i* was then calculated from the following equation where *N*_*k*_ represents the number of openings with a current value *i*_*K*_.

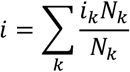

Giant excised membrane patch measurements in inside-out configuration under voltage-clamp conditions were made at room temperature 72-120 h after injection of 50nl channel specific mRNA into Xenopus *laevis* oocytes. Thick-walled borosilicate glass pipettes had resistances of 0.25 - 0.35 MΩ (tip diameter of ∼15 - 30 µm) and were back-filled with extracellular solution containing (in mM): 4 KCl, 116 mM NMDG, 10 HEPES and 3.6 CaCl_2_ (pH was adjusted to 7.4 with KOH/HCl). Bath solution was applied to the cytoplasmic side of the excised giant patches via a gravity flow multi-barrel application system and had the following composition (in mM): 120 KCl, 10 HEPES, 2 EGTA and 1 Pyrophosphate. Currents were acquired with an EPC10 USB amplifier and HEKA PatchMaster 2×91 software. The sampling rate was 10 kHz and the analog filter was set to 3 kHz (−3 dB). Voltage ramp pulses (−80 mV to +80 mV) were applied from a holding potential (V_H_) of -80 mV for 0.8 s with an interpulse interval of 9 s and were analyzed at a given voltage of +40 mV. The relative steady-state levels of inhibition for indicated blocker were fitted with the following Hill equation:

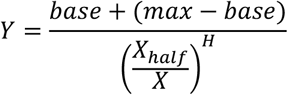

where *base* is the inhibited (zero) current, *max* is maximum current, *x* is the blocker concentration, *X*_*half*_ is the value of concentration for half-maximal occupancy of the blocker binding site and *H* is the Hill coefficient.

Tetrapentylammonium hydrochloride (TPenA-HCl), Bupivacaine hydrochloride monohydrate, A293 (AVE1231), Doxapram hydrochloride and 1,2-Dioctanoyl-*sn*-glycerol (DiC8) were purchased from Sigma-Aldrich. PK-THPP, A1899, Carvedilol and Anandamide (AEA) were purchased from Tocris Bioscience. TASK-1 K_2P_ specific inhibitor BAY1000493 was synthesized by Bayer AG as previously described^23^. All compounds were stored as stock solutions (10 – 100 mM) at -80°C and were diluted in intracellular bath solution to final concentrations prior to each measurement.

### Molecular Dynamics

Simulations were done in GROMACS 2018 using the CHARMM36 force field. Missing atoms in the protein structure (PDB ID: 6RV2) were repaired using SWISS-MODEL, with mutants generated in PyMOL. Each structure was embedded into POPC membranes in independent triplicates and solvated by aqueous NaCl at 150 mM. Temperature and pressure were maintained at 310 K and 1 bar using the velocity-rescaling thermostat and a semi-isotropic Parrinello and Rahman barostat, with coupling constants of 0.1 ps and 1 ps, respectively. Long-range electrostatic interactions were treated using the particle mesh Ewald method. The integration time step was 2 fs. Covalent bonds were constrained through the LINCS algorithm. Pore radii along the channel pathway were determined using the CHAP program^62^.

### Ethics approvals

Ethics committee approvals are available from all participating institutions. The original study^28^ has UK Research Ethics Committee approval (10/H0305/83, granted by the Cambridge South REC, and GEN/284/12 granted by the Republic of Ireland REC).

## Figure Legends

**Table 1. Summary of *KCNK3* variants and phenotypes in nine probands identified with DDSA**.

## Supplementary Figure Legends

**Figure S1:**
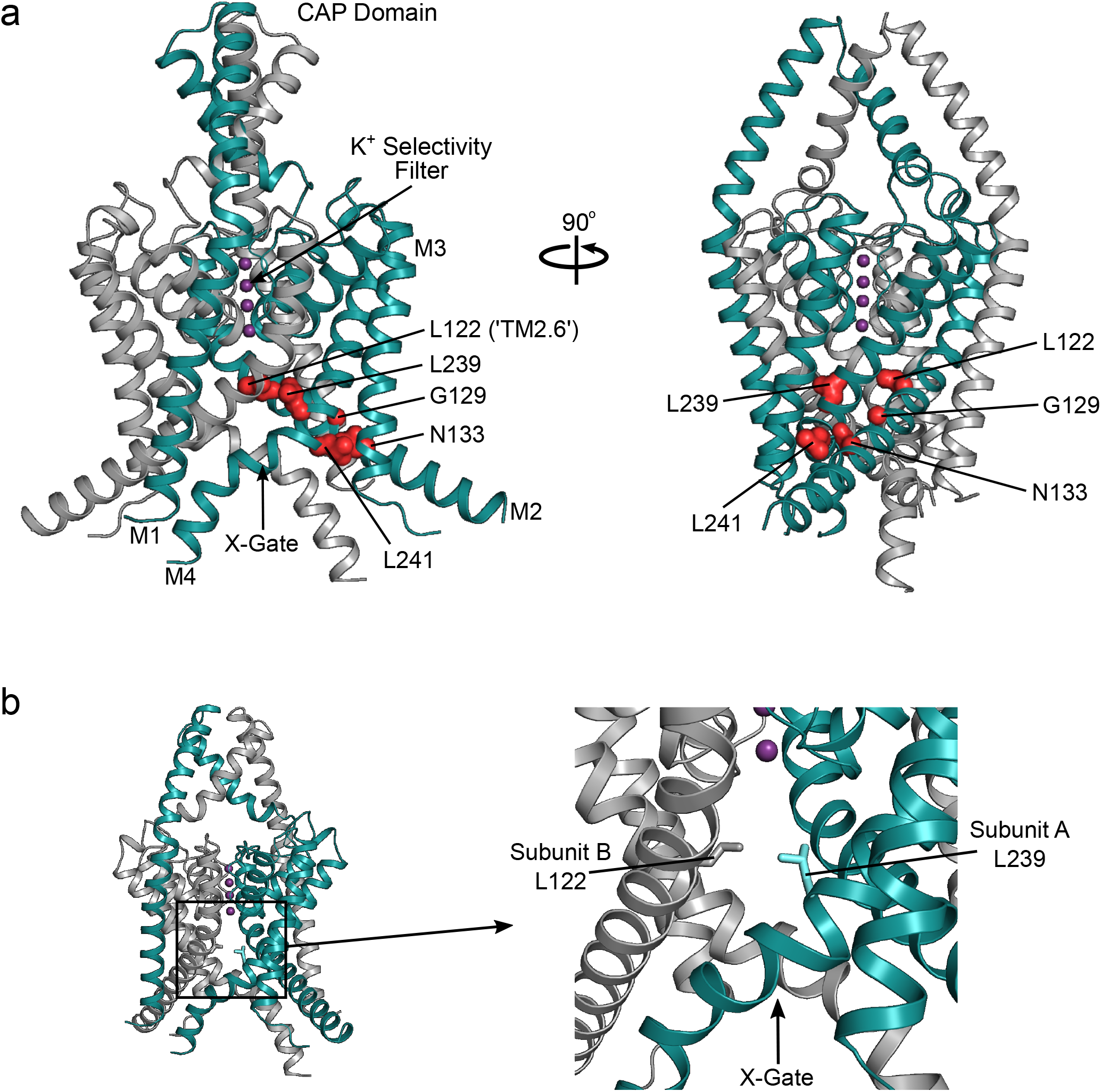
Structure of TASK-1 detailing the location of the DDSA mutations near the X-gate. **a)** Crystal structure of TASK-1 (PDB ID: 6RV2) showing the dimeric nature of the channel. One half of the dimer (subunit A) is highlighted in teal with the DDSA residues shown as spheres and highlighted in red. The selectivity filter, the X-gate, extracellular CAP domain and helices M1-M4 are also marked. The mutated residues all cluster near the X-gate formed by the kink in the M4 helices. **b)** Expanded view of the intracellular X-gate highlighting the inter-subunit proximity of L122 on M2 (subunit B) and L239 on M4 (subunit A). Thus mutation of L122 is likely to disrupt the stability of intersubunit interactions which hold the X-gate closed.

**Figure S2:**
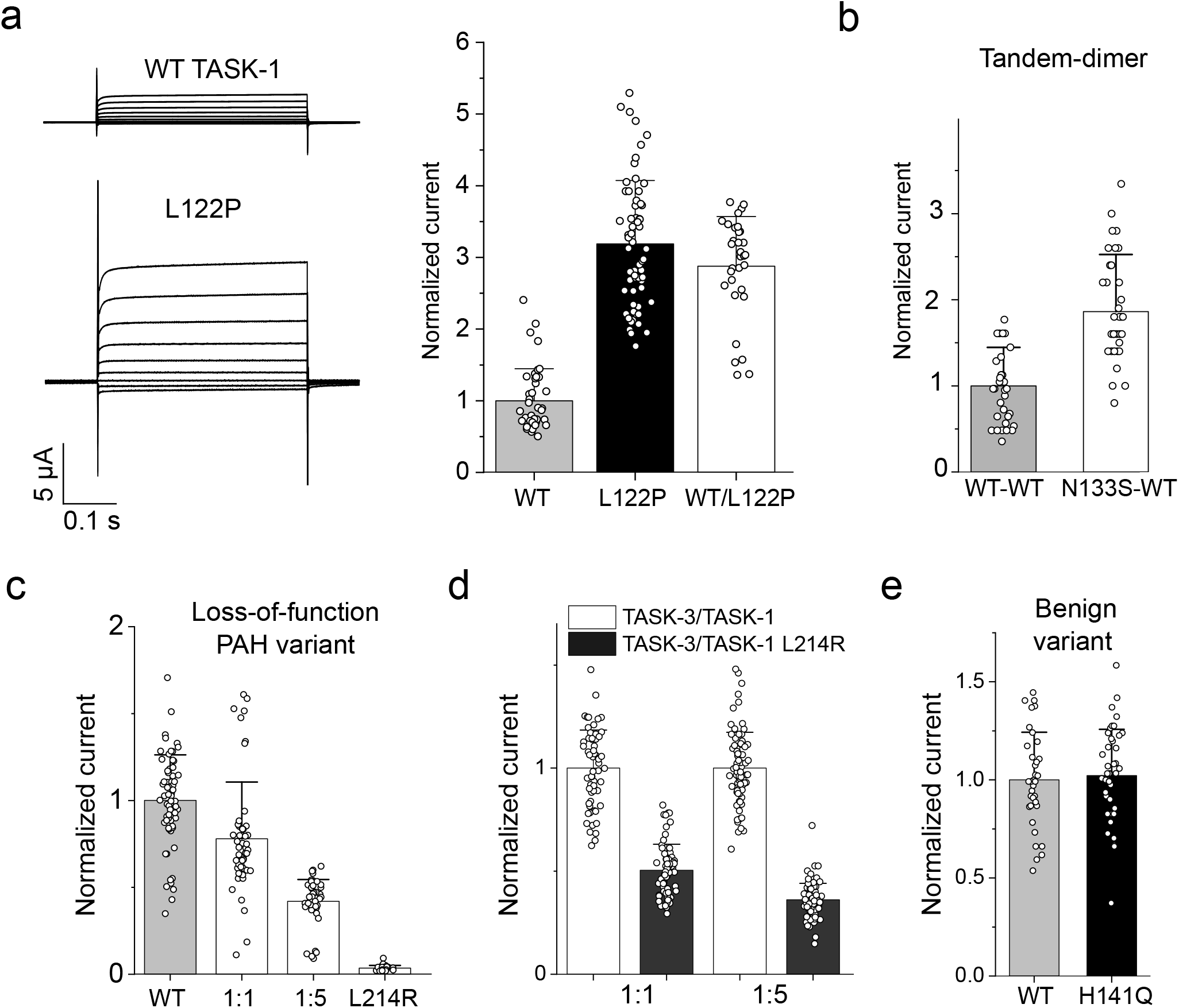
DDSA mutant current levels with PAH and benign variant controls. **a)** Representative currents for WT TASK-1 (n=42) and L122P (n=62), and for 1:1 coexpression of WT TASK-1 with L122P (n=32). **b)** Comparison of tandemly linked WT-WT TASK-1 channels (n=33) and tandemly linked N133S-WT (n=35) channels which limits the stoichiometry of coassembly, currents are normalized to WT-WT currents. **c)** The loss-of-function dominant-negative effect of the L214R PAH mutation in TASK-1 is markedly different to that seen with the gain-of-function DDSA mutations either when coexpressed with WT TASK-1 in different ratios: WT:WT (n=64), WT/L214R 1:1 (n=51), 1:5 (n=48), or L214R alone (n=27), **d)** similar dominant-negative effects of L214R are seen in heteromeric TASK-1/TASK-3 channels: TASK-3/TASK-1 (1:1) (n=63), TASK-3/TASK-1 L241R (1:1) (n=70), TASK-3/TASK-1 (1:5) (n=78), TASK-3/TASK-1 L241R (1:5) (n=85). **d)** Comparison of whole-cell currents at +50 mV for the benign variant in TASK-1, H141Q (n=44) with WT (n=35).

**Figure S3.**
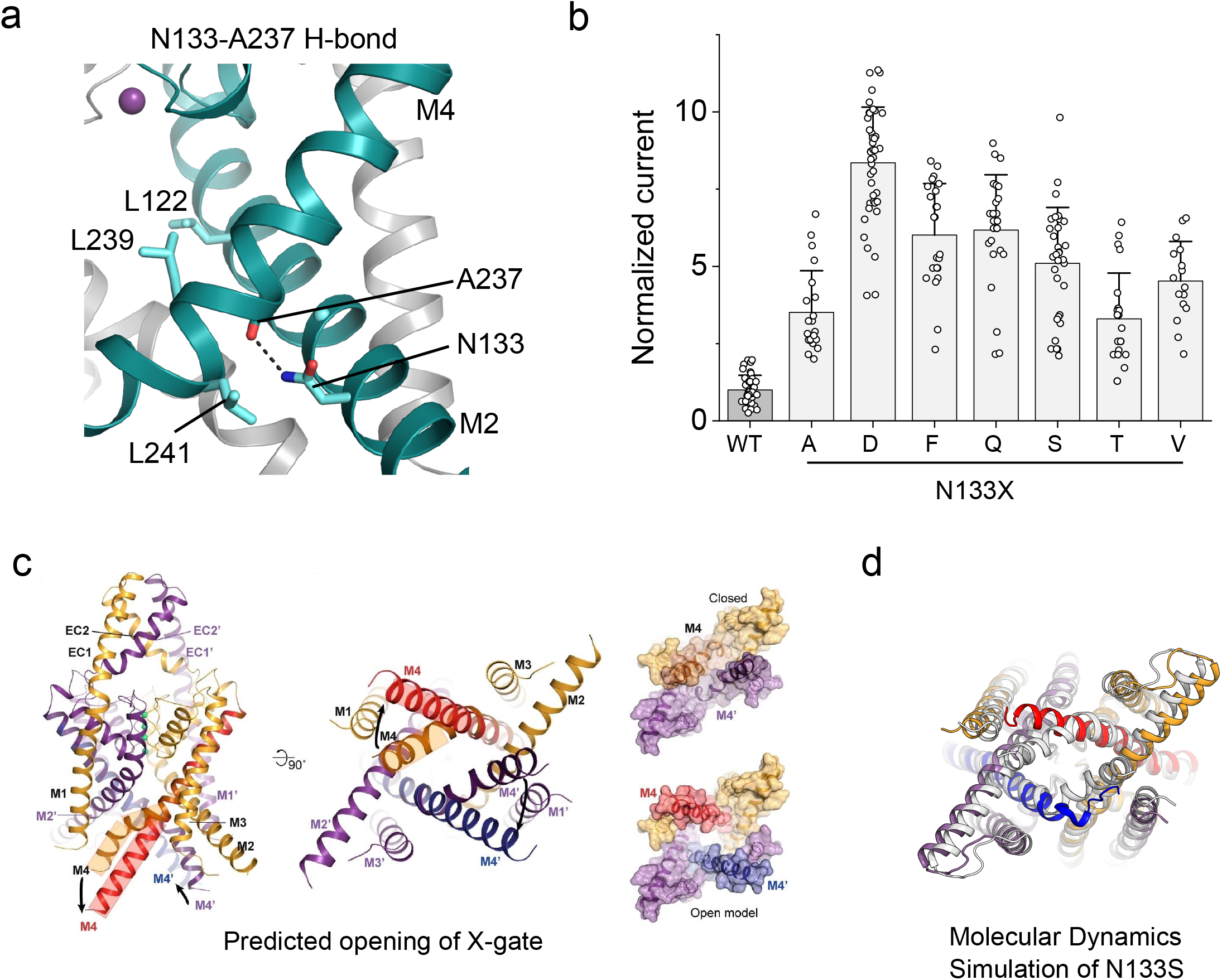
Opening of the X-gate. **a**) Structure of the TASK-1 channel highlighting the intra-subunit hydrogen bond which occurs between N133 on the M2 helix and A237 on the M4 helix. Subunit A is colored teal and the indicated H-bond as a dashed line. **b**) Functional effect of different mutations at position N133: WT (n= 37), N133A (n= 21), N133D (n=42), N133F (n=24), N133Q (n=25) N133S (n=32), N133T n=22, N133V (n=16), currents normalized to WT current at +50 mV, data are presented as mean ± SD. **c**) As previously suggested^23^, for the X-gate to open the M4 helix (colored gold when closed and in red when open) must straighten and rotate for the channel to become open and conductive. d) Molecular dynamics equilibrium simulation of the N133S mutant channel structure embedded within a bilayer indicates a similar movement of the M4 helix (in red) when the X-gate opens; the starting structure is shown in grey.

**Figure S4.**
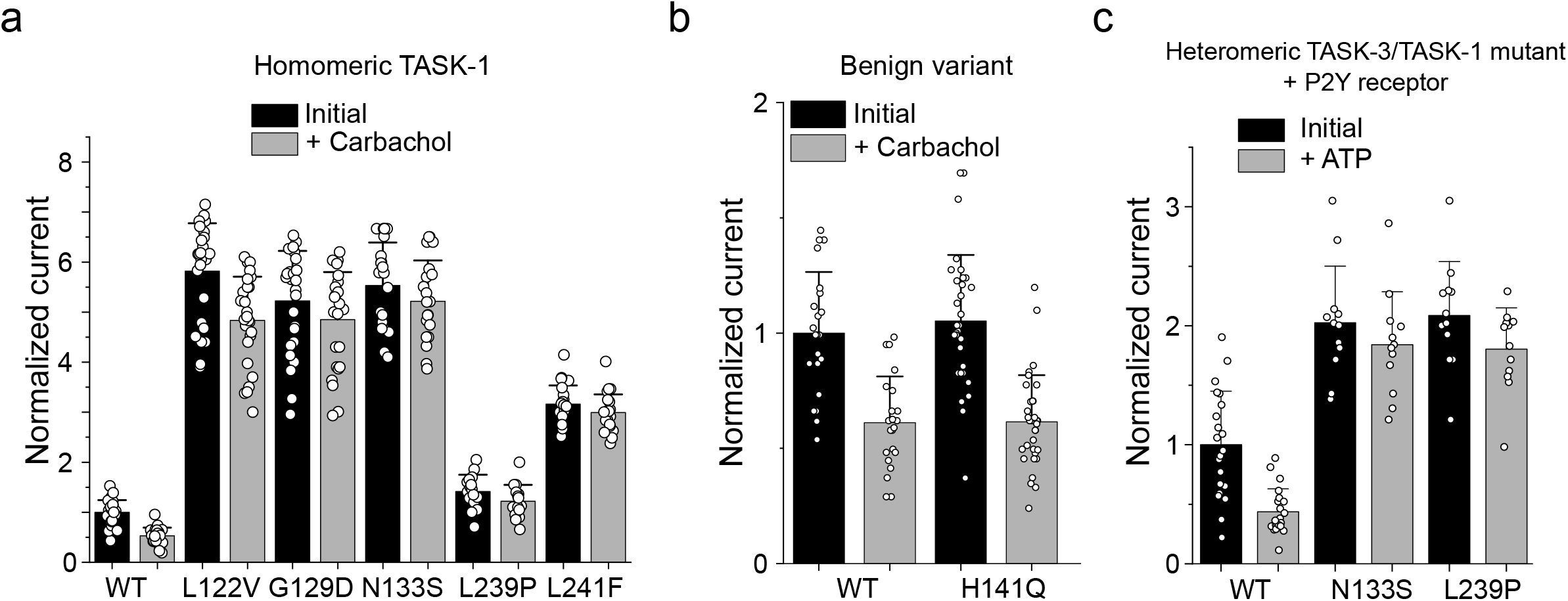
GPCR mediated Inhibition. a) DDSA homomeric channels where both subunits contain the mutation, before and after addition of 10 µM carbachol which normally produces ∼50% inhibition of WT channels: WT TASK-1 (n = 26), L122V (n=27), G129D (n=26), N133S (n=21), L239P (n=16), L241F (n=23), currents are normalized to initial WT currents. Data are presented as mean ± S.D. **b)** The benign mutation does not appear to affect GPCR-mediated inhibition; H141Q (n=33) and TASK-1 (n=22) before and after adding 10 µM carbachol, currents normalized to initial WT current data are presented as mean ± SD. **c)** The N133S mutation in M2 and the L239P mutation in M4 of TASK-1 also severely impair GPCR-mediated inhibition of heteromeric TASK-1/TASK-3 channels.

## References

1. Dempsey, J.A., Veasey, S.C., Morgan, B.J. & O’Donnell, C.P. Pathophysiology of sleep apnea. Physiol Rev 90, 47–112 (2010).

2. Benjafield, A.V. et al. Estimation of the global prevalence and burden of obstructive sleep apnoea: a literature-based analysis. The Lancet Respiratory Medicine 7, 687–698 (2019).

3. Karimi, M., Hedner, J., Habel, H., Nerman, O. & Grote, L. Sleep apnea-related risk of motor vehicle accidents is reduced by continuous positive airway pressure: Swedish Traffic Accident Registry data. Sleep 38, 341–9 (2015).

4. Lyons, M.M., Bhatt, N.Y., Pack, A.I. & Magalang, U.J. Global burden of sleep-disordered breathing and its implications. Respirology 25, 690–702 (2020).

5. Guyenet, P.G. & Bayliss, D.A. Neural Control of Breathing and CO2 Homeostasis. Neuron 87, 946–61 (2015).

6. Niemeyer, M.I., Cid, L.P., Gonzalez, W. & Sepulveda, F.V. Gating, Regulation, and Structure in K2P K+ Channels: In Varietate Concordia? Mol Pharmacol 90, 309–17 (2016).

7. Sepulveda, F.V., Pablo Cid, L., Teulon, J. & Niemeyer, M.I. Molecular aspects of structure, gating, and physiology of pH-sensitive background K2P and Kir K+-transport channels. Physiol Rev 95, 179–217 (2015).

8. Enyedi, P. & Czirjak, G. Molecular background of leak K+ currents: two-pore domain potassium channels. Physiol Rev 90, 559–605 (2010).

9. Mathie, A. Neuronal two-pore-domain potassium channels and their regulation by G protein-coupled receptors. J Physiol 578, 377–85 (2007).

10. Steinberg, E.A., Wafford, K.A., Brickley, S.G., Franks, N.P. & Wisden, W. The role of K2P channels in anaesthesia and sleep. Pflugers Arch 467, 907–16 (2015).

11. Shi, Y. et al. A brainstem peptide system activated at birth protects postnatal breathing. Nature 589, 426–430 (2021).

12. Trapp, S., Aller, M.I., Wisden, W. & Gourine, A.V. A role for TASK-1 (KCNK3) channels in the chemosensory control of breathing. J Neurosci 28, 8844–50 (2008).

13. Talley, E.M., Lei, Q., Sirois, J.E. & Bayliss, D.A. TASK-1, a two-pore domain K+ channel, is modulated by multiple neurotransmitters in motoneurons. Neuron 25, 399–410 (2000).

14. Aller, M.I. & Wisden, W. Changes in expression of some two-pore domain potassium channel genes (KCNK) in selected brain regions of developing mice. Neuroscience 151, 1154–72 (2008).

15. Buckler, K.J. TASK channels in arterial chemoreceptors and their role in oxygen and acid sensing. Pflugers Arch 467, 1013–25 (2015).

16. Olschewski, A. et al. TASK-1 (KCNK3) channels in the lung: from cell biology to clinical implications. Eur Respir J 50(2017).

17. Inoue, M., Matsuoka, H., Harada, K., Mugishima, G. & Kameyama, M. TASK channels: channelopathies, trafficking, and receptor-mediated inhibition. Pflügers Archiv -European Journal of Physiology 472, 911–922 (2020).

18. Hillman, D. A New Pharmacological Treatment to Treat Obstructive Sleep Apnea? Sleep 36, 635–636 (2013).

19. Kiper, A.K. et al. Kv1.5 blockers preferentially inhibit TASK-1 channels: TASK-1 as a target against atrial fibrillation and obstructive sleep apnea? Pflügers Archiv - European Journal of Physiology 467, 1081–1090 (2015).

20. Li, W.-Y. et al. Transient upregulation of TASK-1 expression in the hypoglossal nucleus during chronic intermittent hypoxia is reduced by serotonin 2A receptor antagonist. Journal of Cellular Physiology 234, 17886–17895 (2019).

21. Wang, J., Zhang, C., Li, N., Su, L. & Wang, G. Expression of TASK-1 in brainstem and the occurrence of central sleep apnea in rats. Respiratory Physiology & Neurobiology 161, 23–28 (2008).

22. Cotten, J.F. TASK-1 (KCNK3) and TASK-3 (KCNK9) tandem pore potassium channel antagonists stimulate breathing in isoflurane-anesthetized rats. Anesth Analg 116, 810–6 (2013).

23. Rodstrom, K.E.J. et al. A lower X-gate in TASK channels traps inhibitors within the vestibule. Nature 582, 443–447 (2020).

24. Berg, A.P., Talley, E.M., Manger, J.P. & Bayliss, D.A. Motoneurons express heteromeric TWIK-related acid-sensitive K+ (TASK) channels containing TASK-1 (KCNK3) and TASK-3 (KCNK9) subunits. J Neurosci 24, 6693–702 (2004).

25. Kang, D., Han, J., Talley, E.M., Bayliss, D.A. & Kim, D. Functional expression of TASK-1/TASK-3 heteromers in cerebellar granule cells. J Physiol 554, 64–77 (2004).

26. Ma, L. et al. A novel channelopathy in pulmonary arterial hypertension. N Engl J Med 369, 351–361 (2013).

27. Barel, O. et al. Maternally inherited Birk Barel mental retardation dysmorphism syndrome caused by a mutation in the genomically imprinted potassium channel KCNK9. Am J Hum Genet 83, 193–9 (2008).

28. Kaplanis, J. et al. Evidence for 28 genetic disorders discovered by combining healthcare and research data. Nature 586, 757–762 (2020).

29. Bagriantsev, S.N., Peyronnet, R., Clark, K.A., Honore, E. & Minor, D.L., Jr. Multiple modalities converge on a common gate to control K2P channel function. EMBO J 30, 3594–606 (2011).

30. Piechotta, P.L. et al. The pore structure and gating mechanism of K2P channels. EMBO J 30, 3607–19 (2011).

31. Schewe, M. et al. A pharmacological master key mechanism that unlocks the selectivity filter gate in K(+) channels. Science 363, 875–880 (2019).

32. Talley, E.M. & Bayliss, D.A. Modulation of TASK-1 (Kcnk3) and TASK-3 (Kcnk9) potassium channels: volatile anesthetics and neurotransmitters share a molecular site of action. J Biol Chem 277, 17733–42 (2002).

33. Luethy, A., Boghosian, J.D., Srikantha, R. & Cotten, J.F. Halogenated Ether, Alcohol, and Alkane Anesthetics Activate TASK-3 Tandem Pore Potassium Channels Likely through a Common Mechanism. Mol Pharmacol 91, 620–629 (2017).

34. Ben Soussia, I. et al. Mutation of a single residue promotes gating of vertebrate and invertebrate two-pore domain potassium channels. Nat Commun 10, 787 (2019).

35. Cunningham, K.P. et al. Characterization and regulation of wild-type and mutant TASK-1 two pore domain potassium channels indicated in pulmonary arterial hypertension. J Physiol 597, 1087–1101 (2019).

36. Karczewski, K.J. et al. The mutational constraint spectrum quantified from variation in 141,456 humans. Nature 581, 434–443 (2020).

37. Landrum, M.J. et al. ClinVar: improvements to accessing data. Nucleic Acids Res 48, D835–D844 (2020).

38. Bohnen, M.S. et al. The Impact of Heterozygous KCNK3 Mutations Associated With Pulmonary Arterial Hypertension on Channel Function and Pharmacological Recovery. J Am Heart Assoc 6(2017).

39. Kim, Y., Bang, H. & Kim, D. TBAK-1 and TASK-1, two-pore K(+) channel subunits: kinetic properties and expression in rat heart. Am J Physiol 277, H1669–78 (1999).

40. Ashmole, I. et al. The response of the tandem pore potassium channel TASK-3 (K(2P)9.1) to voltage: gating at the cytoplasmic mouth. J Physiol 587, 4769–83 (2009).

41. Czirjak, G., Petheo, G.L., Spat, A. & Enyedi, P. Inhibition of TASK-1 potassium channel by phospholipase C. Am J Physiol Cell Physiol 281, C700–8 (2001).

42. Srisomboon, Y., Zaidman, N.A., Maniak, P.J., Deachapunya, C. & O’Grady, S.M. P2Y receptor regulation of K2P channels that facilitate K(+) secretion by human mammary epithelial cells. Am J Physiol Cell Physiol 314, C627–C639 (2018).

43. Decher, N., Rinne, S., Bedoya, M., Gonzalez, W. & Kiper, A.K. Molecular Pharmacology of K2P Potassium Channels. Cell Physiol Biochem 55, 87–107 (2021).

44. Wilke, B.U. et al. Diacylglycerol mediates regulation of TASK potassium channels by Gq-coupled receptors. Nature Communications 5, 5540 (2014).

45. Bista, P. et al. The role of two-pore-domain background K(+) (K(2)p) channels in the thalamus. Pflugers Arch 467, 895–905 (2015).

46. Riel, E.B. et al. The versatile regulation of K2P channels by polyanionic lipids of the phosphoinositide (PIP2) and fatty acid metabolism (LC-CoA). bioRxiv, 2021.07.01.450694 (2021).

47. Maingret, F., Patel, A.J., Lazdunski, M. & Honore, E. The endocannabinoid anandamide is a direct and selective blocker of the background K(+) channel TASK-1. EMBO J 20, 47–54 (2001).

48. Wirth, K.J., Steinmeyer, K. & Ruetten, H. Sensitization of Upper Airway Mechanoreceptors as a New Pharmacologic Principle to Treat Obstructive Sleep Apnea: Investigations with AVE0118 in Anesthetized Pigs. Sleep 36, 699–708 (2013).

49. Gurges, P., Liu, H. & Horner, R.L. Modulation of TASK-1/3 channels at the hypoglossal motoneuron pool and effects on tongue motor output and responses to excitatory inputs in vivo: implications for strategies for obstructive sleep apnea pharmacotherapy. Sleep 44(2021).

50. Venken, K.J., Simpson, J.H. & Bellen, H.J. Genetic manipulation of genes and cells in the nervous system of the fruit fly. Neuron 72, 202–30 (2011).

51. Schmidt, C. et al. Upregulation of K(2P)3.1 K+ Current Causes Action Potential Shortening in Patients With Chronic Atrial Fibrillation. Circulation 132, 82–92 (2015).

52. Deciphering Developmental Disorders, S. Large-scale discovery of novel genetic causes of developmental disorders. Nature 519, 223–8 (2015).

53. Deciphering Developmental Disorders, S. Prevalence and architecture of de novo mutations in developmental disorders. Nature 542, 433–438 (2017).

54. McKenna, A. et al. The Genome Analysis Toolkit: a MapReduce framework for analyzing next-generation DNA sequencing data. Genome Res 20, 1297–303 (2010).

55. McLaren, W. et al. The Ensembl Variant Effect Predictor. Genome Biology 17, 122 (2016).

56. Ramu, A. et al. DeNovoGear: de novo indel and point mutation discovery and phasing. Nat Methods 10, 985–7 (2013).

57. Wright, C.F. et al. Genetic diagnosis of developmental disorders in the DDD study: a scalable analysis of genome-wide research data. Lancet 385, 1305–14 (2015).

58. Retterer, K. et al. Clinical application of whole-exome sequencing across clinical indications. Genet Med 18, 696–704 (2016).

59. Di Donato, N. et al. Mutations in EXOSC2 are associated with a novel syndrome characterised by retinitis pigmentosa, progressive hearing loss, premature ageing, short stature, mild intellectual disability and distinctive gestalt. J Med Genet 53, 419–25 (2016).

60. Thorvaldsdóttir, H., Robinson, J.T. & Mesirov, J.P. Integrative Genomics Viewer (IGV): high-performance genomics data visualization and exploration. Briefings in Bioinformatics 14, 178–192 (2013).

61. Aryal, P., Abd-Wahab, F., Bucci, G., Sansom, M.S. & Tucker, S.J. A hydrophobic barrier deep within the inner pore of the TWIK-1 K2P potassium channel. Nat Commun 5, 4377 (2014).

62. Klesse, G., Rao, S., Sansom, M.S.P. & Tucker, S.J. CHAP: A Versatile Tool for the Structural and Functional Annotation of Ion Channel Pores. J Mol Biol 431, 3353–3365 (2019).

